# “Crochet … a little hook to improve attention?”

**DOI:** 10.1101/2022.12.17.22283453

**Authors:** Davide Rossi Sebastiano, Cristina Muscio, Dunja Duran, Deborah Bonfoco, Sara Dotta, Paola Anversa, Pietro Tiraboschi, Elisa Visani

**Affiliations:** Neurophysiology Unit, Fondazione IRCCS Istituto Neurologico Carlo Besta, 20133 Milan, Italy; Azienda Socio-Sanitaria Territoriale- Bergamo Ovest, 24047 Bergamo, Italy; Epilepsy Unit, Fondazione IRCCS Istituto Neurologico Carlo Besta, 20133 Milan, Italy; Unit of Neurology V and Neuropathology, Fondazione-IRCCS-Istituto Neurologico Carlo Besta, 20133 Milan, Italy

**Author notes:** corresponding author **Corresponding author:** Davide Rossi Sebastiano, MD, PhD, Neurophysiology Unit, Fondazione-IRCCS-Istituto Neurologico Carlo Besta, 20133 Milan, Italy. Mail. **Author contributions Davide Rossi Sebastiano:** conceptualization (lead), supervision, writing-original draft; **Cristina Muscio:** conceptualization (supporting); project administration; **Dunja Duran:** formal analysis (supporting), software (equal); **Deborah Bonfoco:** data-curation (equal); **Sara Dotta:** data-curation (equal); **Paola Anversa:** data-curation (equal); **Pietro Tiraboschi:** resource, funding acquisition, writing-review & editing; Elisa Visani: formal analysis (lead), software (equal), visualization.

## Abstract

In this work we compared the short-term effects of crochet on the performance in the well-known Attention Network Test and on the global cortical functioning networks revealed by magnetoencephalography between a group of crocheters and a sex and age-matched control group.

Our data revealed that crochet is associated with an increase of the alerting and the orienting networks even after a brief, single work session and that this behavioural effect seems to have a counterpart in the modification seen in the global functional connectivity of the brain, where an increased speed of the information exchange between different brain areas have been seen. Moreover, we discuss the hypothesis that these effects on attentional networks are dissimilar from those determined by meditation, where an improvement in the executive control was previously demonstrated as the main effect.

Our results provided for the first time that crochet is associated with an increase in the attentional networks, and namely in alerting and orienting networks, paving the way for the use of textile-related arts in the neurorehabilitation, possibly in combination with meditation, considering that the two practices promote complementary effects on the attentional networks.

**Highlights:**

- Crocheting positively affects attention, improving alerting and orienting
- Crocheting speeds up the information exchange between different brain areas
- Crocheting and meditation promote different effects on the attentional networks

## 1. Introduction

One of the most important cornerstones of the occupational therapy is the close relationship between a goal-directed action requiring the physical involvement and the mental health (Gallagher, Muldoon, & Pettigrew, 2015). In this line, two studies focused on the effects of crocheting (Burns & Van Der Meer, 2021) and knitting (Riley, Corkhill, & Morris, 2013) suggested the link between the textile-related arts and wellbeing.

The creativity stimulated by the conception of an object and the repetitiveness of the actions have been advocated as the main reason to explain relaxation, stress relief and a sense of accomplishment promoted by knitting and crocheting (Burns & Van Der Meer, 2021; Riley et al., 2013; Turney J., 2009). The knitting and crocheting fans strongly believe in the several positive effects that their occupation may provide for mental health and happiness. Previous studies revealed that textile–related arts have beyond the well-known, beneficial influence on health as stress-relieving activities, an impact in promoting the attention and in facilitating the learning of new skills. (Brooks, Ta, Townsend, & Backman, 2019; Burns & Van Der Meer, 2021; Riley et al., 2013). In recent years, all these benefits contributed to reviving an old-fashioned craft as a portable, “cool” hobby, potentially useful even for therapeutic and/or social purposes (Gjernes, T., 2017; Guitard et al., 2018), even though, to date, how and whether crocheting and knitting determine modifications in the cortical neural networks has not yet been explored.

As for textile-related arts, the meditation increases wellbeing, reducing psychological stress and improving mental health (Goyal et al., 2014); moreover, it is also associated with the increase of attention and performance (Lippelt, Hommel, & Colzato, 2014; Lutz, Slagter, Dunne, & Davidson 2008) and the global functioning of cortical networks (Jang et al., 2011; Luders, Toga, Lepore, & Gaser, 2009; Tang, Holzel, & Posner, 2015; Tang, Tang, & Posner, 2014; van Lutteveld et al., 2017; Xue,);

The current number of studies on crocheting is very low, especially when compared with the number of works focused on meditation. We hypothesize that the execution of brief, single-session of crochet could improve the functioning of attentional networks, similarly to the meditation.

The Attention Network Test (ANT) and subsequent variants can evaluate all the three attentional components (i.e. alerting, orienting and executive control, Petersen & Posner, 2012; Posner & Petersen, 1990, Callejas, Lupianez, Funes, & Tudela, 2005).

ANT was profitably used in patients with Attention-Deficit/Hyperactivity Disorder, Alzheimer disease, Multiple Sclerosis, and schizofrenia(Dotare, et al., 2020; Fernandez-Duque & Black, 2006; Ishigami, Fisk, Wojtowicz, & Klein, 2013; Orellana, Slachevsky, & Peña, 2012), in healthy children (Ishigami & Klein, 2010), in younger (Callejas et al., 2005, Fan, McCandliss, Sommer, Raz, & Posner, 2002) and older adults (Ishigami et al., 2016), proving its validity and reliability in studying the modifications of the attentional networks.

Our study aimed to identify the effect of crocheting on the functional brain networks, comparing the modifications of the cortical activity detected by Magnetoencephalography (MEG) with the behavioural scores obtained at the ANT after a single-session of crochet in a population of experienced crocheters, in comparison with controls.

## 2. Methods

### 2.1 Study Population

Forty-one people experts in crochet (CRO, 40 females, age ranging from 33 to 65, mean = 52.2±7.6 years) and twelve subjects with no experience in crochet and/or knitting (control group, CTR, 12 females, age ranging from 32 to 73, mean: 49.3±11.3 years) were enrolled, from January 2021 to July 2022, to perform a behavioural-MEG study. All participants were adult (>18 years), right-handed, according to the Edinburgh Handedness Inventory (Oldfield, R. C., 1971) and had normal or corrected-to-normal vision.

All the volunteers of CRO group usually perform crochet and knitting; we included in the study only people who crocheted at least 1 hour/day for at least 5 days/week along the last two months before the study initiation. For both groups, exclusion criteria were the presence of concomitant neurological disease and the current use of drugs affecting the central nervous system. This study was carried out in accordance with the ethical standards of the Declaration of Helsinki and its later amendments, and it was approved by the Ethical Committee of Fondazione-IRCCS-Istituto Neurologico “Carlo Besta” of Milan, Italy (approval number: 76/2020; date of approval: October 7th, 2020). Participants gave their written informed consent before the inclusion in the study.

### 2.2 Protocol and experimental design

The protocol included five phases: MEG acquisition in resting condition (a), execution of ANT task (b), 20 minutes of awake resting (CTR group) or crocheting (CRO group) (c), MEG acquisition in resting condition (d), execution of ANT task (e). In the following parts of the manuscript, reference will be made to T0 for (a) and (b) and to T1 for (d) and (e) phases, respectively. The experimental design is shown in Figure 1.A.

**Figure 1.**
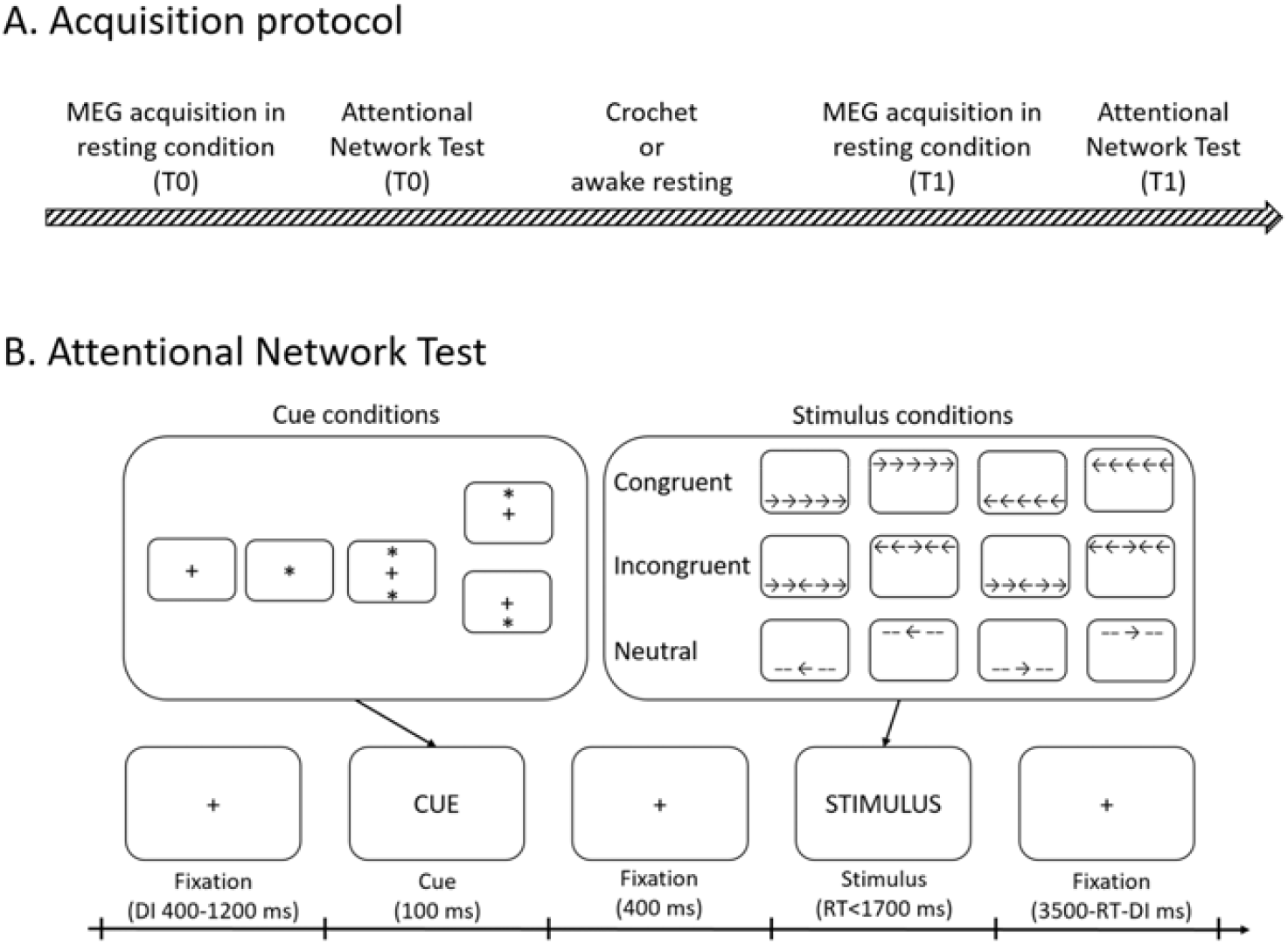
Legend: Experimental design and the behavioral test

The whole protocol took place inside the shielded room of the MEG for a total duration of about one hour.

#### 2.2.1 Acquisition in resting condition

The acquisition in resting condition consisted in 3 minutes with eyes open and 3 minutes with eyes closed. Participants were asked to remain still as much as possible, and avoid blinking and falling asleep. The acquisition in resting condition was repeated twice, before (T0) and after (T1) the crochet work or rest.

#### 2.2.2 Behavioural task

The ANT task primarily described by Fan et al. (2002) was used, implementing the stimuli on Stim2 software (Compumedics-Neuroscan; Compumedics Limited, Abbotsford, Victoria, Australia; Neuroscan, Charlotte, NC, USA). Responses were detected through two button devices (Current Designs Inc., Philadelphia, PA, USA), one in each hand.

During ANT task, cues (asterisks) and stimuli (arrows) were presented on a screen positioned in front of the subject. The task requires participants to determine whether a central arrow (target) points left or right. Each trial started with a fixation cross displayed for a time randomly ranging from 400 and 1200 ms. A cue was then presented for 100 ms and followed by a fixation cross with 400 ms of duration. The cue can be one of the following types: no cue, center cue, double cue (one at the top and one at the bottom of the screen) and spatial cue (one at the top or at the bottom of the screen). Subsequently, the stimulus was presented for 1700 ms. The stimuli were associated to three conditions: neutral, congruent and incongruent. The target was flanked by two lines on both sides in the neutral condition or by two arrows in the other two conditions. The arrows pointed toward the same direction as the target arrow in the congruent condition or in the opposing direction in the incongruent condition. The target always appeared at the location of the asterisk in the spatial-cue condition, and the target positions varied (above or below) randomly in other conditions. After participants made a response, the stimulus disappeared immediately and a fixation cross was set again (Figure 1.B).

The ANT was repeated twice, before (T0) and after (T1) the crochet; the two blocks of ANT were randomized: each of them included 288 trials and lasted about 17 minutes. A practice block of 20 trials was conducted before the experiment to allow participants to become familiar with the task and the recording environment.

For all the other participants, the reaction time (RT) of each stimulus was determined as the time length between the onset of the target stimulus and the response detected by pressing the button device. The alertness network RT was calculated by subtracting the cue from the no cue RT. The orienting network RT was calculated as the difference between center cue and spatial cue RT. The executive control network RT was calculated by subtracting the congruent from the incongruent RT. Data deriving from individuals who made more than 10% of errors of the total in the behavioural test were excluded from the subsequent analysis

#### 2.2.3 Crochet and rest activity

After the first execution of the first ANT, all the CRO participants performed a crochet activity which consisted of reproducing a simple pattern (granny square) to complete a hexagonal tile of about 10 cm diagonal, for a total duration of about 20 minutes. Participants were asked to work in the usual manner and speed. All the equipment for crochet, included the wool balls, and the wooden, MEG-compatible hooks were provided by the no-profit organization Gomitolorosa ONLUS.

The subjects of the CTR group were asked to stay in a relaxed and quiet resting condition for a total duration of 20 minutes. To ensure that the subject did not fall asleep, the subject’s MEG trace was monitored also during this phase and, and in case of signs of drowsiness, the subject was prompted via intercom.

### 2.3 MEG

#### 2.3.1 MEG data acquisition and pre-processing

MEG signals were acquired using a 306-channel whole head MEG system (Triux, Elekta Oy, Helsinki, Finland) with a sampling frequency of 1 kHz. Bipolar electro-oculographic (EOG) and electrocardiographic signals (ECG) were also acquired. The participant’s head position inside the MEG helmet was continuously monitored by five Head Position Identification (HPI) coils located on the scalp. The locations of HPI, together with three anatomical landmarks (nasion, right and left preauriculars), and additional scalp points were digitized before the recording by means of a 3D digitizer (FASTRAK, Polhemus, Colchester, VT, Usa).

The raw MEG data were pre-processed off-line using the spatio-temporal signal space separation method (Taulu & Simola, 2006) implemented in the Maxfilter 2.2 (Elekta Neuromag Oy, Helsinki, Finland) to subtract external interference and correct for head movements using HPI signals.

Physiological artifacts were removed through use of Signal-Space Projectors (Gross et al., 2001) using EOG and ECG signals to mark eye movements or blinks and heartbeat, respectively. Finally, data were band-pass filtered at 1–100 Hz. To characterize instrument and environmental noise for source modelling, noise covariance matrix was estimated using 2 minutes of empty-room recordings collected just before each session.

Individual MEG data were co-registered with a template MRI (MNI/ICBM152, Fonov et al., 2011) using digitized scalp points. Then, MEG forward model was calculated using the overlapping-spheres approach. Based on the Desikan-Killiany atlas, regions of interest (ROIs) were defined in the subject’s co-registered MRI. The time-series of neuronal activity were reconstructed by projecting sensor signals to source space using dynamic Statistical Parametric Mapping (dSPM) method (Dale et al., 2000). To obtain a single time series per ROI, a mean of all vertices of each ROI was calculated after correcting for an opposite sign source direction. The analysis was performed using Brainstorm software (Tadel et al., 2011).

#### 2.3.2 Functional connectivity analysis

For each participant of the CRO and CTR group, 90 consecutive seconds of MEG signal in the open-eye resting condition before and after crochet performance (CRO) or relaxing (CTR) were selected and then segmented into 45 non-overlapping epochs with a duration of 2 s each. From these epochs, different indices of Functional Connectivity (FC) were calculated in the classical theta (4–8 Hz), alpha (8–13 Hz) and beta (13–30 Hz) frequency bands, while frequencies <4 Hz and >30 Hz were excluded from analysis.

The weighted Phase Lag Index (wPLI, Stam & Reijneveld, 2007) was estimated for each frequency bands in each cortical ROI and then as mean overall ROIs (global FC). The results were averaged over the epochs for each subject, thus obtaining a FC matrix for each frequency band.

Network topology was examined using Minimum Spanning Tree (MST) graphs derived from the functional connectivity matrices (Stam et al., 2014). The MST graph is a sub-network derived from a weighted network including the highest weights possible without forming any loop. Consequently, the resulting graph has the same number of links, allowing comparison across groups or conditions without running the risk of bias due to differences in edge density. MST is proposed to reflect the functional core of the network (Van Mieghem & Magdalena, 2005).

To study the network integration, the following global connectivity measures were extracted from MST graph: maximum Betweenness Centrality (BC), Diameter, and Tree Hierarchy (TH). BC measures the central network organization in terms of importance of the most central node. High values of diameter indicate a decreased global efficiency. TH characterize the hypothesized optimal topology of brain network organization, where information is transferred between brain regions in the fewest possible steps (i.e., star-like topology), while preventing information overload of central brain regions. A schematic depiction of the MSTs calculation pipeline is shown in Figure 2.

**Figure 2.**
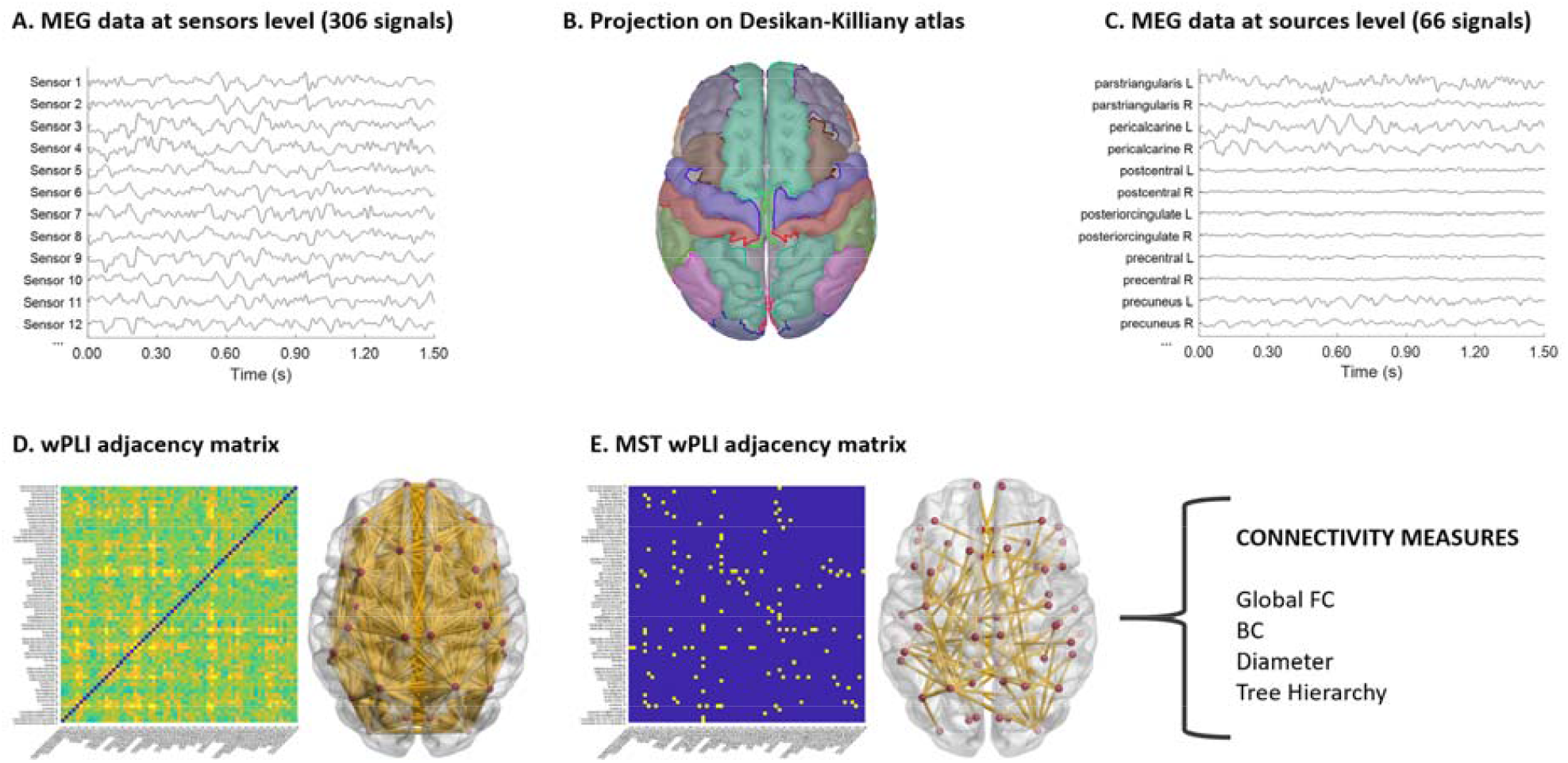
Legend: Schematic depiction of the Minimum Spanning Trees calculation pipeline

Analyses were performed using custom Matlab (MATLAB 2021a, MathWorks, Inc., Natick, MA, USA) based on Fieldtrip (Oostenveld, Fries, Maris, & Schoffelen, 2011) and BCT (Rubinov & Sporns, 2010) toolboxes.

### 2.4 Statistical analysis

Accuracy and RTs were compared using repeated measures ANOVA (rmANOVA) with GROUP (CRO and CTR) as between factor and TIME (T0 and T1) as within factor. rmANOVA was applied also to compare cue type (center, spatial and no cue) and stimulus type (congruent, incongruent, neutral) as within factor. The same analysis was applied for network scores global FC and connectivity indexes for each band separately.

The sphericity assumption was evaluated using Mauchley’s test and the Greenhouse-Geisser degree of freedom correction was applied when appropriate. Post-hoc tests were performed by means of paired or independent t-test with Bonferroni correction for multiple comparison. Statistical significance was set at p<0.05. Values are expressed as mean ± standard error of the mean. All statistical analyses were performed with SPSS (IBM Statistic, version 28.0).

## 3. Results

### 3.1 Behavioural task

All volunteers adequately carried out the protocol. One subject of the CRO group was excluded from all analyses due to technical problems during the acquisition, whereas three CRO subjects and one CTR subject were excluded from the analyses because they made too many errors in behavioural task (error rate 11.5%, 12.8%, 10.8% and 12.8%, respectively). Therefore, 36 subjects from the CRO group and 11 for the CTR group were included in the analysis

For the remaining subjects in the first test (T0) the mean error rate was 6.9% and 8.7% for CRO and CTR respectively, while in the second test (T1) was 3.0% and 4.8%.

For both groups, the accuracy increased between the first and second test (CTR: 96.86±0.49% vs. 98.2±0.69%; CRO: 96.91±0.47% vs. 98.61±0.28%). rmANOVA showed a significant main effect of TIME on accuracy and global RTs (F(1,46)=12.94, p<0.001 and F(1,46)=18.83, p<0.001, respectively). Post-hoc test revealed that the accuracy significantly increase in both groups (CTR: t(10)=-4.06, p=0.002; CRO: t(36)=-3.84, p<0.001), whereas the mean RTs significantly decrease (CTR: t(10)=2.17, p=0.027; CRO: t(36=4.85, p<0.001). On average, in both groups, the RTs decreased in all condition (Table 1).

**Table 1.** Reaction time (RT) of attentional test. Bold text indicates significant difference between T0 (first test) and T1 (second test) after correction for multiple comparison. Italic text indicates results not surviving the correction (trend).

|  |  | Center cue<br>(ms) | No cue<br>(ms) | Spatial cue<br>(ms) | Mean RT<br>(ms) | T0 vs T1 |
| --- | --- | --- | --- | --- | --- | --- |
| **CRO group** |  |  |  |  |  |  |
| Congruent | T0 | 503.2±12.3 | 517.8±13.0 | 486.7±11.2 | **503.3±11.7** | t(36)=4.97, |
|  | T1 | 481.0±11.4 | 481.9±12.2 | 451.1±10.4 | **473.0±10.8** | p<0.001 |
| Neutral | T0 | 509.5±11.8 | 505.5±11.0 | 476.5±12.0 | **495.8±11.3** | t(36)=3.56, |
|  | T1 | 477.6±11.6 | 493.5±11.6 | 457.1±12.3 | **474.5±11.2** | p=0.001 |
| Incongruent | T0 | 552.2±13.0 | 551.3±10.9 | 516.8±12.7 | **540.5±11.8** | t(36)=5.33, |
|  | T1 | 521.7±12.6 | 517.6±11.4 | 476.6±11.8 | **506.2±11.6** | p<0.001 |
| Mean | T0 | **520.8±12.0** | **524.6±11.4** | **493.2±11.6** | **513.1±11.4** |  |
|  | T1 | **493.4±11.5** | **497.6±11.4** | **461.1±11.3** | **484.6±11.1** |  |
|  | T0 vs | t(36)=4.55, | t(36)=4.24, | t(36)=5.19, | t(36)=4.85, |  |
|  | T1 | p<0.001 | p<0.001 | p<0.001 | p<0.001 |  |
| **CTR group** |  |  |  |  |  |  |
| Congruent | T0 | 520.2±26.7 | 535.5±23.5 | 493.9±29.2 | 513.9±24.7 | t(10)=2.07, |
|  | T1 | 496.2±24.2 | 499.3±24.2 | 462.7±24.0 | 485.2±22.5 | p=0.065 |
| Neutral | T0 | 508.3±26.3 | 513.2±22.1 | 480.5±24.0 | 499.8±23.5 | t(10)=1.68, |
|  | T1 | 483.9±22.4 | 496.9±24.2 | 464.8±24.0 | 481.0±22.6 | p=0.123 |
| Incongruent | T0 | 559.8±25.1 | 561.4±23.1 | 511.0±26.0 | 543.6±24.5 | t(10)=2.60, |
|  | T1 | 521.4±23.2 | 526.8±23.0 | 484.4±26.8 | 512.1±22.9 | p=0.026 |
| Mean | T0 | 528.5±25.6 | 536.6±22.7 | 495.1±25.9 | 518.9±24.0 |  |
|  | T1 | 500.2±23.5 | 507.6±22.3 | 470.7±24.7 | 492.7±22.4 |  |
|  | T0 vs | t(10)=2.29, | t(10)=2.03, | t(10)=2.06, | t(10)=2.17, |  |
|  | T1 | p=0.045 | p=0.070 | p=0.066 | p=0.055 |  |

For the different cue types, rmANOVA showed a significant effect of TIME (F(1,46)=19.02, p<0.001) and cue types (F(1.5,69.48)=48.63, p<0.001). Comparing separately the group between first and second test, RTs decreased in both groups, but significantly only in CRO group (Table 1). For the different stimulus type, rmANOVA showed a significant effect of TIME (F(1,46)=18.97, p<0.001), stimulus types (F(1.25,57.31)=95.06, p<0.001) and a significant interaction between them (F(1.73,79.37)=929.35, p<0.001). Comparing first and second test, post-hoc tests revealed RTs decrease for related to all stimulus type for both groups, but the differences were significant only in CRO group (Table 1).

rmANOVA showed a significant effect of time for alertness and orienting networks (F(1,46)=6.72, p=0.013 and F(1,46)=5.41, p=0.024). Post-hoc tests revealed that the difference was significant for the CRO group (ART: t(36)=-2.79, p=0.004; ORT: t(36)=2.97, p=0.003), but not for the CTR group (ART: t(10)=-1.59, p=0.072; ORT: t(10)=1.32, p=0.109, Table 2).

**Table 2.**
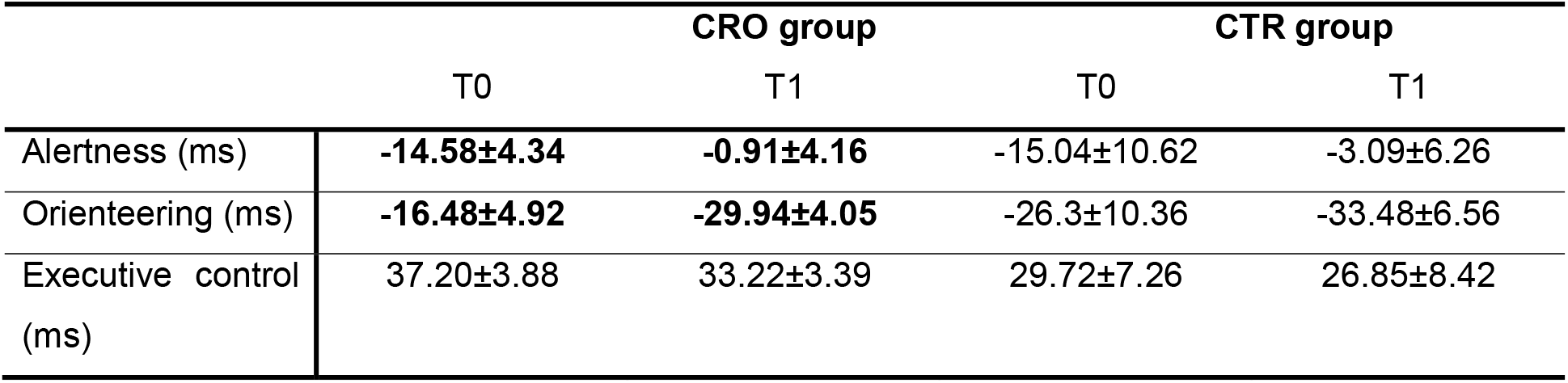
Alertness, orienteering and executive control networks results. Bold indicates significant difference between T0 (first test) and T1 (second test).

|  |  | **CRO group** |  | **CTR group** |  |
| --- | --- | --- | --- | --- | --- |
|  |  | T0 | T1 | T0 | T1 |
| Alertness (ms) |  | **-14.58±4.34** | **-0.91±4.16** | -15.04±10.62 | -3.09±6.26 |
| Orienteering (ms) |  | **-16.48±4.92** | **-29.94±4.05** | -26.3±10.36 | -33.48±6.56 |
| Executive control<br>(ms) |  | 37.20±3.88 | 33.22±3.39 | 29.72±7.26 | 26.85±8.42 |

### 3.2 Functional connectivity

rmANOVA on global FC in alpha band showed significant main effect of TIME (F(1,46)=5.06, p=0.029) and a strong trend toward significance of between groups difference (F(1,46)=3.00, p=0.090). Post-hoc tests revealed that the global FC was significantly higher at T1 with respect to T0 for the CRO group (t(36)=-3.79, p=0.003), but not for the CTR group (t(10)=-0.917, p=0.381). When comparing connectivity value at T1 between groups, a trend for higher values in the CRO than CRT group was found (t(46)=-1.75, p=0.087). No other differences were found.

rmANOVA on BC values in beta band showed a significant effect of TIME (F(1,46)=4.91, p=0.032). Post-hoc revealed that BC values increase at T1 with respect to T0 in the CRO (t(36)=-2.40, p=0.022) but not in the CRT group (t(10)=-1.40, p=0.191). rmANOVA on Tree Hierarchy in alpha band measures showed a robust trend toward significance of between groups factor (F(1,46)=3.85, p=0.056). Post-hoc-test showed that the values increased almost significantly at T1 only in the CRO group (t(36)=-1.77, p=0.085) whereas they didn’t change in the CRT group (t(10)=0.20, p=0.845).

Results of FC are shown and plotted in Table 3 and Figure 3, respectively.

**Table 3.** Comparison of minimum spanning tree measures during resting condition. Bold indicates significant difference between T0 (first recording) and T1 (second recording).

|  |  | **CTR**<br>**T0** | **CTR**<br>**T1** | **CRO**<br>**T0** | **CRO**<br>**T1** |
| --- | --- | --- | --- | --- | --- |
| Theta | wPLI | 0.36±0.00 | 0.36±0.00 | 0.36±0.00 | 0.36±0.00 |
|  | BC | 0.69±0.02 | 0.69±0.02 | 0.68±0.01 | 0.70±0.01 |
|  | Diameter | 15.3±0.82 | 16.6±0.94 | 15.65±0.50 | 15.35±0.52 |
|  | Tree Hierarchy | 0.37±0.00 | 0.37±0.01 | 0.38±0.01 | 0.37±0.01 |
| Alpha | wPLI | 0.27±0.00 | 0.28±0.00 | **0.28±0.00** | **0.29±0.01** |
|  | BC | 0.71±0.02 | 0.75±0.03 | 0.71±0.01 | 0.69±0.01 |
|  | Diameter | 14.3±0.91 | 12.4±0.75 | 13.38±0.57 | 13.40±0.46 |
|  | Tree Hierarchy | 0.39±0.01 | 0.39±0.01 | **0.40±0.01** | **0.42±0.01** |
| Beta | wPLI | 0.15±0.00 | 0.16±0.00 | 0.16±0.00 | 0.16±0.00 |
|  | BC | 0.69±0.01 | 0.72±0.03 | **0.68±0.00** | **0.72±0.01** |
|  | Diameter | 14.10±1.10 | 14.8±0.91 | 14.70±0.58 | 15.03±0.53 |
|  | Tree Hierarchy | 0.39±0.02 | 0.37±0.02 | 0.40±0.01 | 0.38±0.01 |

Legend. wPLI, weighted Phase Lag Index; BC, betweenness centrality.

**Figure 3.**
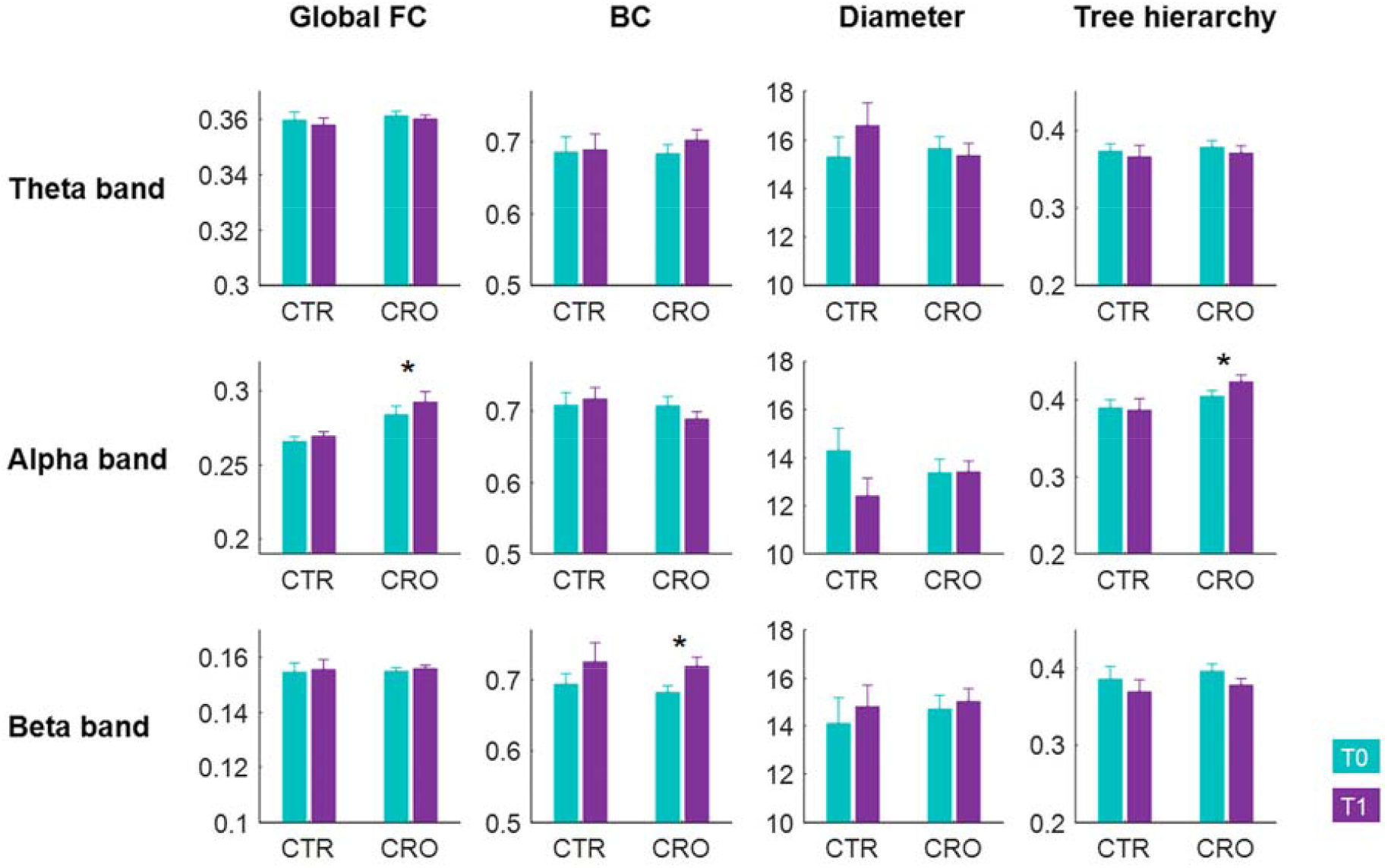
Legend: Boxplots of the global connectivity measures for theta, alpha and beta bands at T0 (cyan) and T1 (purple). *=statistically significant at p<0.05

## 4. Discussion

### 4.0 Crocheting improve alerting and orienting and determine change in the functional connectivity

We found that crochet seems to improve alerting and orienting networks because a significant effect of time (T0 vs T1) on RT (for all cue and stimulus type) and alertness and orienting networks (but not for executive control) was found for the CRO but not for the control group. As these scores usually remain constant after repeated sessions even in an older population (Ishigami & Klein, 2010), the improvements in alerting and orienting scores can be attributed to the crochet performance.

Many works demonstrated that attention is composed by three separate anatomical and functional systems, i.e., alerting, orienting and executive control (conflict) networks, which, albeit independent, cooperate and influence each other to produce an adaptive and performing behavior (Fan et al., 2002; Fernandez-Duque & Posner, 1997; Petersen and Posner, 2012; Posner and Petersen, 1990; Raz & Buhle, 2006). Alerting is the ability to enhance and maintain readiness in preparation for an impending and imminent stimulus, while orienting selectively allocates attentional resources to specific information among multiple ones (Fernandez-Duque & Posner, 1997; Petersen and Posner, 2012; Posner and Petersen, 1990; Raz & Buhle, 2006). From a functional point of view, alerting and orienting systems are so largely interdependent that some stimuli are able to produce an unconscious activation on the alerting and orienting networks, despite their failure to reach awareness (Lu et al., 2012; Peterson & Gibson, 2011). In an ecological context, probably alerting and orienting networks cooperate in influencing the speed of reacting to environmental events; in a laboratory setting, there are further evidence that the global activation provided by the alerting network can make faster the orienting process to the salient stimulus/stimuli (Callejas et al., 2005).

Data of FC are coherent with the behavioural ones: in resting condition, we observed a TH in alpha band and BC in beta band significantly higher at T1 with respect to T0 for CRO group, but not for CTR group. TH measures the optimal topology of brain network organization, where information is transferred between different cortical areas in the fewest possible steps (and therefore in the shortest possible time), while preventing information overload of central brain regions (Tewarie, van Dellen, Hillebrand, & Stam, 2015; van Dellen et al., 2014). Again, BC indicates the number of shortest paths passing through a definite node and it is related to the central network as expressed in terms of importance of the most pivotal among them (Tewarie et al., 2015; van Dellen et al., 2014). Since increased TH and BC values usually tend to correspond to an increased network integration, our results showed that a single session of crochet could promote the alerting and orienting networks functioning probably increasing the information exchange between different brain areas.

### 4.1 Similarities and differences of crocheting and meditation on the attentional networks

Many works demonstrated that meditation can increase performance (Lippelt, Hommel, & Colzato, 2014; Lutz, Slagter, Dunne, & Davidson 2008) and even promote neuroplasticity inducing both functional and even long-lasting (structural) changes in the brain networks (Luders, Toga, Lepore, & Gaser, 2009; Tang, Holzel, & Posner, 2015). Local and global efficiency of cortical networks are reported to increase in meditators both during meditation (Jang et al., 2011) and in a resting state (van Lutteveld et al., 2017; Xue, Tang, Tang, & Posner, 2014). Moreover, previous works demonstrated that an enhancement of the performances could be achieved even after a single-session meditation (Chan, Immink, & Lushington, 2017).

In light of all these similarities of the effects reported for meditation and textile-related arts, the hypothesis behind our study was that crocheting can act in promoting the functioning of attentional networks in an analogous way to meditation. We especially expected that crochet would produce effects like those of the Focused Attention Meditation (FAM), a meditation in which practitioners must continuously focus the concentration on a chosen object or internal physiological condition (breathing) to prevent mind wandering (Lutz et al., 2008). Surprisingly, our findings showed unexpected differences between crocheting-related and meditation-related effects on attentional networks which must be accounted for. Previous work argued that both FAM and Open-Monitoring Meditation (OMM, a meditation which does not involve a deliberate selection of stimuli) determine significant improvement in executive control network, but not in alerting nor in orienting ones, nor in global RT in ANT (Ainsworth, Eddershaw, Meron, Baldwin, & Garner, 2013; Jha, Krompinger, & Baime, 2007). Again, Tang et al., 2007, showed an analogous improvement in the executive control networks after integrative body–mind training (a kind of meditation akin to OMM). On the contrary, in our study crocheting improved global RT, alerting, and orienting networks, but not executive control.

From a neurophysiological point of view, so far it has been proved that meditation promotes an increase in the brain network integration since, compared with novice meditators, experienced meditators showed significantly higher BC and LF and significantly lower diameter and average eccentricity in alpha band (van Lutterveld et al., 2017). Moreover, Xue et al. (2014) showed larger clustering coefficient, local and global efficiency, and shorter average path length in theta band after short-term meditation. Finally, increased cortical frontal midline theta activity were present during internally guided states of FAM, when compared to periods of rest with mind wandering or spontaneous thought (Aftanas & Golocheikine, 2001; Brandmeyer & Delorme, 2018), in line with observations that frontal midline theta activity is involved in sustaining internalized attention, especially for OMM (Cavanagh & Frank, 2014; Travis & Shear, 2010).

On the contrary, we found an increase in TH in alpha band and in BC in beta band for the CRO group (see results 3.1 and discussion, 4.2). Thus, the behavioral and neurophysiological data converge in hypothesizing a different mechanism in promoting attention between meditation and crochet. Meditation usually refers to a wide range of mental activities which emphasize attention-based, regulatory training regimes (Lutz, Jha, Dunne, & Saron, 2015), the goal of which is to maintain a monitoring state, remaining attentive to any arising stimuli or experience, without selecting, judging, or focusing on any particular thing (Kabat-Zinn, 2009). Hence, meditation could be considered as a top-down process which can support a mindfulness of the cognitive control, by allowing for fast and volitional shifts of alternate attentional profiles due to a better executive control functioning which results in a more effective decision making. On the other hand, crocheting involves coordinated and asymmetric movement of fingers and wrists, in turn complicated by the need for continuous and prompt attentional shifts from the general plot to the manual gesture, in order to carry out the original project (Riley et al., 2013). In this view, crocheting could be considered as an intensive bimanual rehabilitative exercise able to promote a bottom-up, unconscious training of the alerting and orienting networks, as a side effect of its intrinsic needs to achieved the pursued goal.

### 4.3 Limitations

Previous studies demonstrated that meditation was associated with increased FC between the default mode network, the dorsal attention network, and the right prefrontal cortex (Lippelt et al., 2014). They especially highlighted the effect of meditation on the functioning of anterior cingulate cortex, indicating its higher network efficiency (Xue et al., 2014) and its role in detecting mind-wandering and feeding it back to executive control networks, in order to re-focus the attention (Carter & van Veen, 2007; Lippelt et al., 2014). These evidences clarified the effects of meditation on the functioning of definite brain areas, thus providing a coherent link between the experimental data and the theoretical framework of the executive control function (Petersen and Posner, 2012). In our study we focused the analysis on global indices of the FC, while we did not explore the effect at a single brain area level. Further studies are needed in order to explore the involvement of the single areas.

Moreover, we measured cortical magnetic activity before and after, but not during the crochet performance, since the possible presence of movement-related artefacts on MEG signals. Further studies based on high-resolution EEG recording could bridge this gap. We chose to use the MEG to minimize the discomfort of the volunteers who underwent the measure and the overall duration of the experiment.

## 5. Conclusion

In this study, we investigated for the first time the short-term effects of crochet on the performance in a well-known attentional test and on the global cortical functioning networks revealed by MEG, in a group of crocheters, compared with a sex and age-matched control group.

Our study showed that crocheting is positively associated with a boosting of the alerting and the orienting functions even after a brief, single work session and that this behavioural effect has a counterpart in the modification seen in the global FC of the brain, where an increased speed of the information exchange between different brain areas was demonstrated.

## Data Availability

All data produced in the present study are available upon reasonable request to the authors

## Acknowledgments

We want to thank Gomitolorosa ONLUS and the Italian Ministry of Health which supported our study.

Gomitolorosa ONLUS (https://www.gomitolorosa.org) is a non-profit association that encourages the recovery of native wool and is even committed to projects for the increase of the well-being of patients engaged in medical therapies by means of the promotion of textile-related arts among them. Our thanks go to all the enrolled volunteers and especially to dr. Alberto Costa and dr. Ivana Appolloni who supported us in the execution of the study.

